# Intraprocedural continuous saline infusion lines significantly reduce the incidence of acute kidney injury during endovascular procedures for stroke and myocardial infarction: evidence from a systematic review and meta-regression

**DOI:** 10.1101/2023.10.05.23296627

**Authors:** Gianluca De Rubeis, Simone Zilahi De Gyurgyokai, Sebastiano Fabiano, Luca Bertaccini, Andrea Wlderk, Francesca Romana Pezzella, Sabrina Anticoli, Giuseppe Biondi Zoccai, Francesco Versaci, Luca Saba, Enrico Pampana

## Abstract

**Background:** Contrast media used in mechanical therapies for stroke and myocardial infarction represent a significant cause of acute kidney injury (AKI) in acute medical scenarios. Although the continuous saline infusion line (CSIL) is a standard procedure to prevent thrombus formation within the catheter during neurovascular interventions of mechanical thrombectomy (MT), it is not utilized in percutaneous coronary interventions (PCI).

**Material and methods:** A systematic review of the incidence of AKI after MT for stroke treatment was performed. These data were compared with those reported in the literature regarding the incidence of AKI after PCI for acute myocardial infarction. A random-effect model meta-regression was performed to explore the effects of CSIL on AKI incidence, using clinical details as covariates.

**Results:** A total of 18 and 33 studies on MT and PCI were included, respectively, with 69,464 patients (30,138 [43.4%] for MT and 39,326 [56.6%] for PCI). The mean age was 63.6 years ±5.8 with male 66.6% ±12.8. Chronic kidney disease ranged 2.0%–50.3%. Diabetes prevalence spanned 11.1% to 53.0%. Smoking status had a prevalence of 7.5%–72.0%. Incidence of AKI proved highly variable (I^2^=98%, Cochrane’s Q 2985), and appeared significantly lower in the MT subgroup than in the PCI subgroups (respectively 8.3% [95% confidence interval: 4.7%–11.9%] vs 14.7 [12.6%– 16.8%], p<0.05). Meta-regression showed that CSIL was significantly associated with a decreased incidence of AKI (OR=0.93 [1.001–1.16]; p=0.03).

**Conclusion:** Implementation of CSIL during endovascular procedures in acute settings was associated with a significant decrease in the risk of AKI, and its safety should be routinely considered in such interventions.

**Condensate abstract:** Acute kidney injury (AKI) has an incidence rate of 21.6% in the acute setting. The occurrence of AKI during acute myocardial infarction (AMI) increases the chance of death with an OR of 12.52 (95% CI 9.29–16.86). This study explored the effect of continuous saline infusion line (CSIL) on the incidence of AKI by comparing mechanical thrombectomy and percutaneous coronary intervention. Meta-regression showed that CSIL was significantly associated with a decreased incidence of AKI (OR=0.93 [1.001–1.16]; p=0.03). By implementing CSIL as a standard operative procedure in PCI, 1 out of 9 deaths could be prevented during AMI treated with PCI.

**What is Known:**

- Contrast media induces acute kidney injury.
- Acute kidney injury increases mortality in ischemic stroke and acute myocardial infarction
- Saline infusion has been used to prevent and treat acute kidney injury.
- Continue saline infusion is used in neurovascular intervention for preventing thrombus formation

**What the Study Adds:**

- Continue saline infusion during percutaneous arterial treatment reduces the incidence of acute kidney injury

**Conflict of Interest statement:** GBZ: Amarin, Balmed, Cardionovum, Crannmedical, Endocore Lab, Eukon, Guidotti, Innovheart, Meditrial, Microport, Opsens Medical, Terumo, and Translumina, outside the present work. The remaining authors have no COI

The manuscript complies with all instructions to authors

The authorship requirements have been met and the final draft was approved by all authors

**A list of each author’s contributions:** Conceptualization GDR and SZG; methodology GDR and LS, software GDR; validation formal analysis GDR, SA, EP, SF, GBZ, FRP, FV; investigation GDR And MA; resources LB and AW; data curation GDR and SZG; writing—original draft preparation GDR; writing—review and editing LS, EP, SF, SA, FRP, GBZ, FV; visualization GDR; supervision LS, EP, SF, SA, FRP, GBZ, FV;

The manuscript has not been published elsewhere and is not under consideration by another journal

The paper adherences to ethical guidelines and indicate ethical approvals (IRB) and use of informed consent, as appropriate. IRB approvals was not necessary being the paper a systematic review and meta-analysis

**Reporting checklist:** PRISMA

*TOC category:* Coronary intervention

*Classification:* Clinical

## Introduction

Acute kidney injury (AKI) has an incidence of 21.6% (95% confidence interval [95% CI], 19.3 to 24.1) in the acute setting(1) It has been widely demonstrated that iodine contrast administration may increase the risk of developing AKI due to transient endothelium-dependent vasodilation, and vasoconstriction potentially critically reduces renal blood flow and induces renal ischemia (2,3). Although there are no clear differences between the routes of administration of contrast media in inducing AKI (e.g., intravenous or intra-arterial), a lower threshold value of the glomerular filtration rate was chosen for intra-arterial administration(4). However, some studies have emphasized, for the risk of developing AKI, patient comorbidities, including diabetes and preexisting chronic kidney disease(2,4). However, all these studies presented several confounding factors for the case mix included(4).

Patients with acute conditions were more prone to develop AKI (incidence 21.6% [95% confidence interval [95% CI], 19.3 to 24.1]). Ischemic stroke (IS) and acute myocardial infarction (AMI) have several clinical similarities, including pathophysiology, acute onset, prognosis, and effective treatment (5). Furthermore, both diseases can be treated endovascularly using mechanical thrombectomy (MT) for IS or percutaneous coronary intervention (PCI) for AMI(6,7). Considering that both clinical presentations have the same setting, including acute disease and intra-arterial contrast administration, these patients are prone to develop AKI(6,7).

To avoid distal embolization, neurointerventions are performed under continuous saline infusion (CSI) (8), which is not mandatory for PCI(9). During MT, a continuous saline infusion line (CSIL) is connected to all catheters used during the procedure. Although it is difficult to evaluate the amount of saline administered during MT, it can be estimated that approximately 361.8 to 1085.4 ml depending on the number of catheters used (e.g., 2 co-axial catheter 2 saline infusion) and time of the procedure(10,11). Interestingly, fluid expansion is one of the main strategies used to prevent and treat AKI (12). Furthermore, the amount of saline infused during MT is similar to that recommended by the European Society of Urogenital Radiology (ESUR) to prevent and treat AKI(4,13).

This systematic review, meta-analysis, and meta-regression aimed to evaluate the impact of CSIL on the incidence of AKI during endovascular procedures for the treatment of acute diseases. Therefore, we compared the incidence of AKI after MT with that after PCI for AMI.

## Material and methods

The study protocol is available upon reasonable request from the corresponding author. In addition, the data used for the systematic review were publicly available. The study was drafted according to the PRISMA guidelines(14). The review protocol was registered in PROSPERO (ID 433497). The search strategy for the MT subgroup was discussed in the following paragraph. The results were compared with those of Lun et al.(15) on the incidence of AKI in PCI on 1.2 millions of patients in 2021.

### Mechanical thrombectomy subgroup: search strategy and selection criteria

We conducted a systematic search of electronic databases, including PubMed, OVID (“MEDLINE® and Epub Ahead of Print, In-Process, In-Data-Review & Other Non-Indexed Citations, Daily and Versions”), and the Cochrane Library, from their inception to march-2023. We used a combination of MeSH terms and keywords related to “acute kidney injury,” “mechanical thrombectomy,” “contrast induce nephropathy”, and “stroke” by using different Boolean operators (AND or OR). The PICO question was: “Is continuous saline infusion during trans-arterial percutaneous interventions (MT and PCI) effective in reducing acute kidney injury in the acute setting (IS and AMI)?. The exclusion criteria were non-English literature and fewer than 10 patients, as the study sample size. We excluded studies that did not report the incidence of AKI or clinical outcomes. A complete list of these articles is presented in **Table 1**.

**Table 1.**
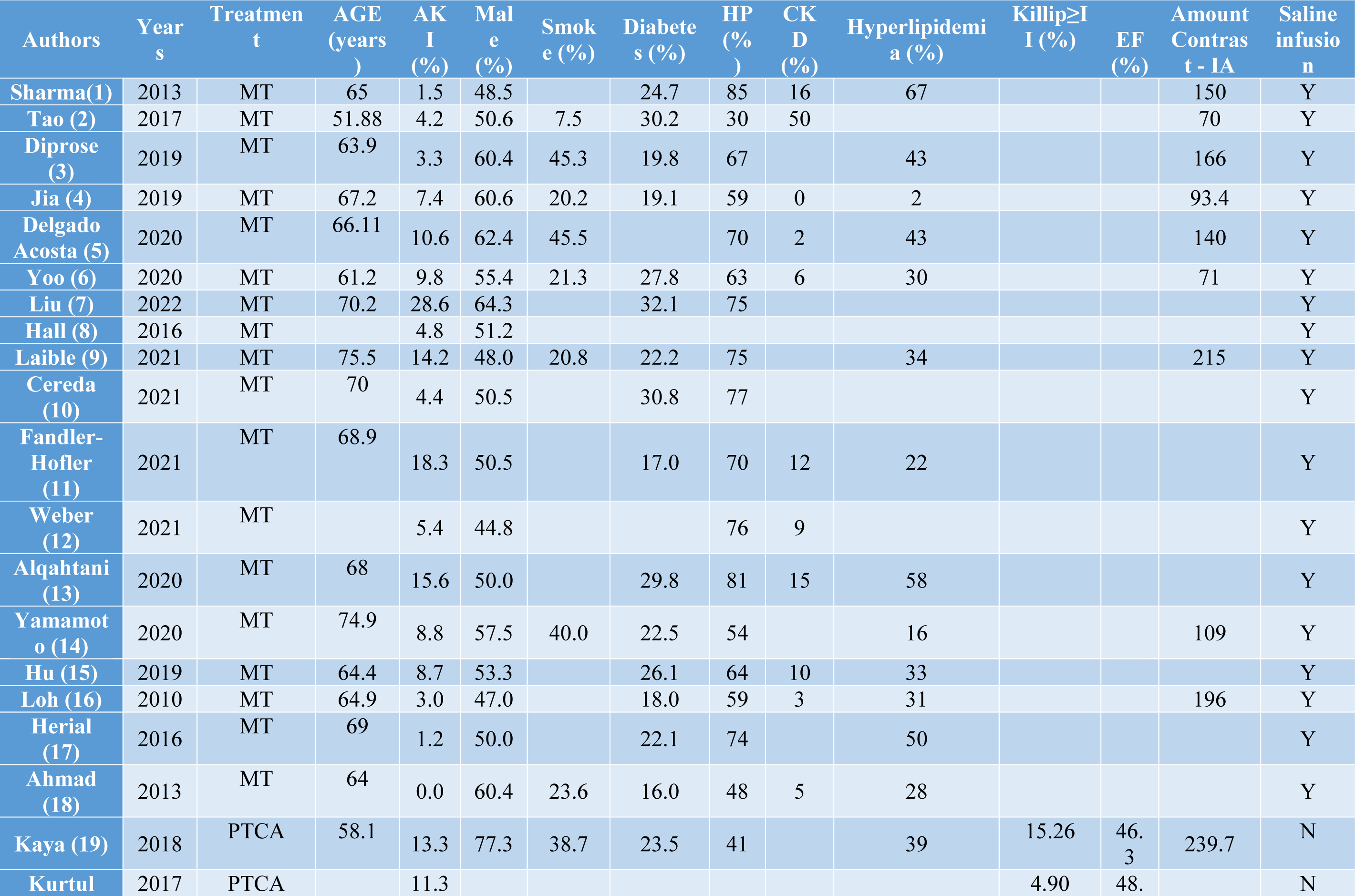
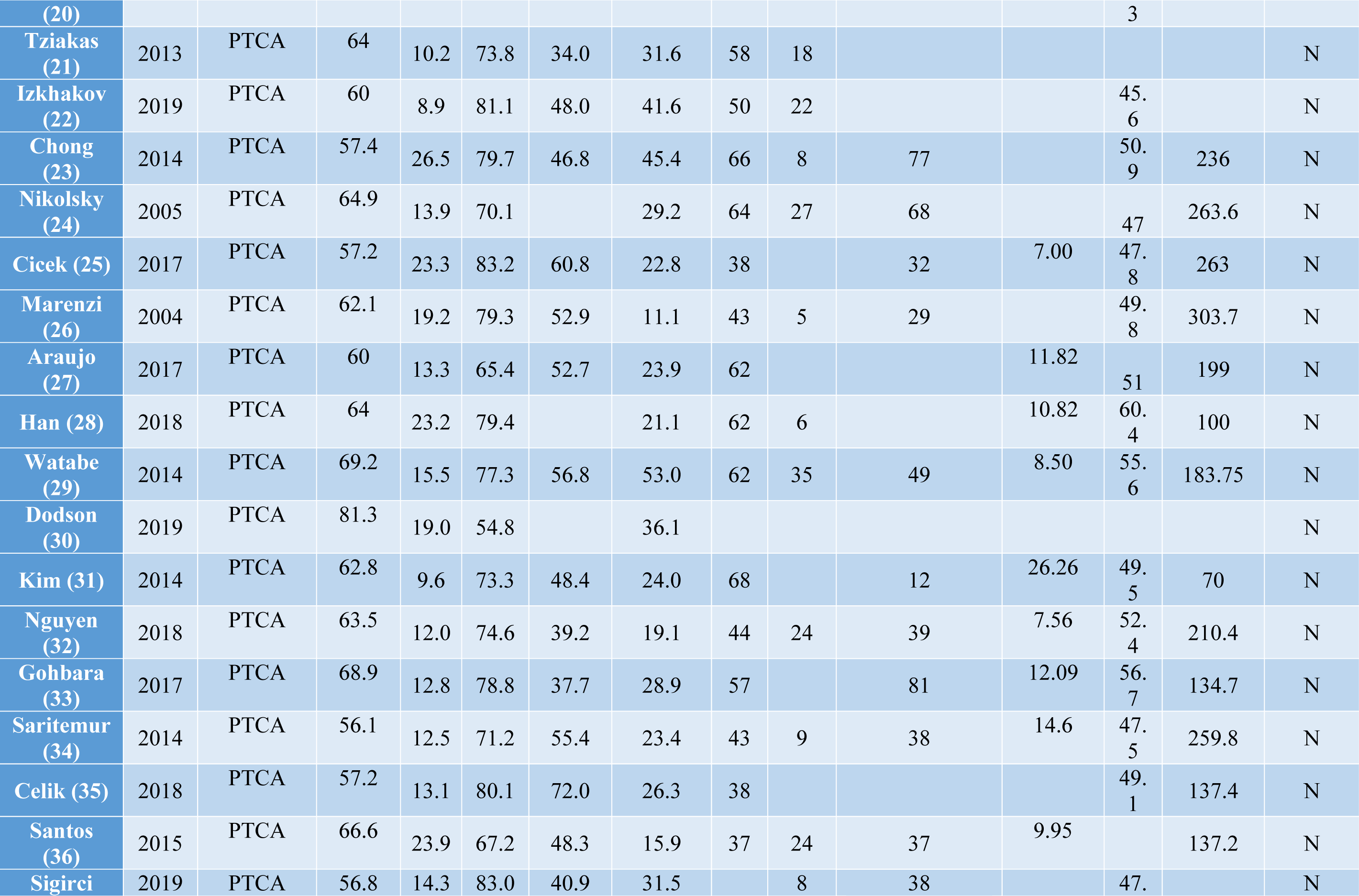
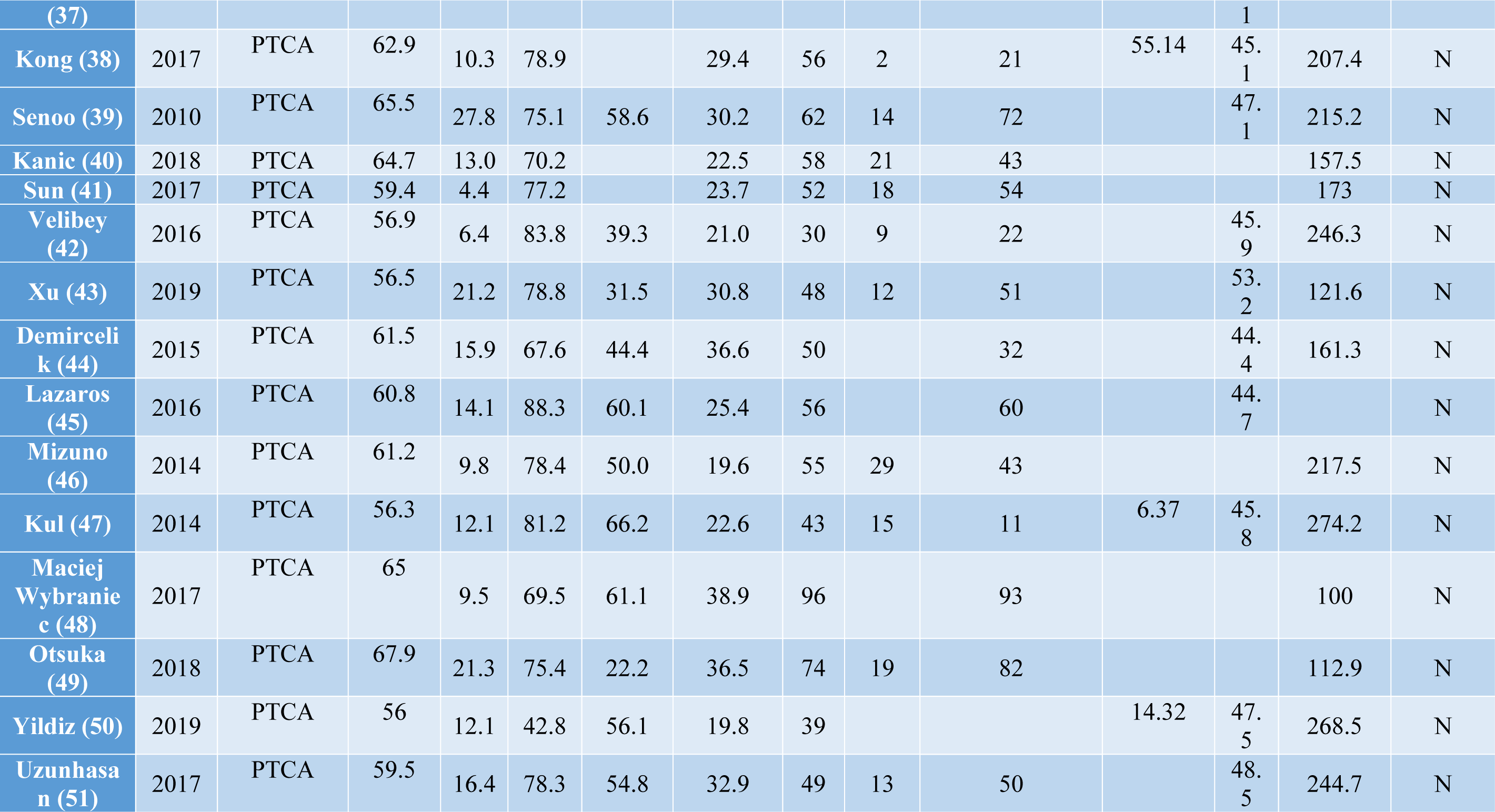
Studies details. MT: mechanical thrombectomy; PTCA: percutaneous coronary angioplasty; AKI acute kidney injury OR: odds ratio; CKD chronic kidney disease; HP: hypertension; IA: inter-arterial; Y: yes; N: no

### Percutaneous coronary interventions

All studies (n = 133) included in the systematic review by Lun et al.(15) were retrieved, and the study characteristics, patient demographics, comorbidities, and AKI incidence were reported. Furthermore, only studies in the acute setting were included to reduce the confounding factors (33/133 [24.8%]). This was performed to ensure a proper comparison between acute ischemic stroke and acute myocardial infarction. The research was not re-performed because it is highly consistent with the large sample size of the study (>1.2 million patients) and the updating of the data (published in 2021).

### Data extraction, quality assessment and statistical analysis

For the MT subgroup, an automatic deduplication tool was used for the Ovid. Subsequently, Rayyan (https://www.rayyan.ai) was used for duplication screening(16). Two reviewers independently (two neurointerventional radiologist [five and three years of experience]) extracted data on study characteristics (e.g., study design, sample size, and year of publication), patient demographics (e.g., age, sex, and comorbidities) and AKI incidence (see **Table 1**). Disagreements were resolved through consensus. The risk of bias was assessed using the Methodological Index for Non-randomized Studies (MINORS) for observational studies. (**Table S1**). The body of evidence was evaluated with Grading of Recommendations, Assessment, Development, and Evaluations (GRADE)(17) (**Table S2**)

Data were reported using the mean, median, standard deviation, and confidence interval 95% (CI95%), according to the distribution. Publication bias was assessed by using funnel plots. I^2 and Cochrane’s Q were used for measuring heterogeneity.

A subgroup meta-analysis using random effect models was performed to compare the incidence of AKI between MT and PCI. To investigate the potential reasons for this discrepancy, meta-regression was performed using a random effect model in both univariate and multivariate analyses. Meta-regression was used to evaluate the effect of CSIL independently from other comorbidities and demographic details, including age, gender, smoking status, diabetes, prior chronic kidney disease (CKD) and amount of contrast media administrated. To evaluate the potential source of heterogeneity, the analysis was also performed without the study by Alqahtani et al.(18) because the population of the study in the MT subgroup was more than 70% of the total (73.6%). Sensitivity analysis was performed using leaving-one-out meta-analysis.

All analyses were conducted using R-Studio (R-project http://www.R-project.org) and OpenMeta version 12.11.14 (http://www.cebm.brown.edu/openmeta/index.html) were used as statistical software.

## Results

### Search results

Our systematic search yielded 3317 articles. A total of 57/3317 (1.7%) were automatically deleted by the Rayyan AI for duplications. A total of 97/3260 (3.0%) studies were manually excluded because of duplications. 3136/3163 (99.1%) were excluded after reviewing the title and abstract. After retrieving the full text, 9/27 (33.3%) papers were excluded. The final population for stroke treatment included 18 studies (**Figure 1**).

**Figure 1.**
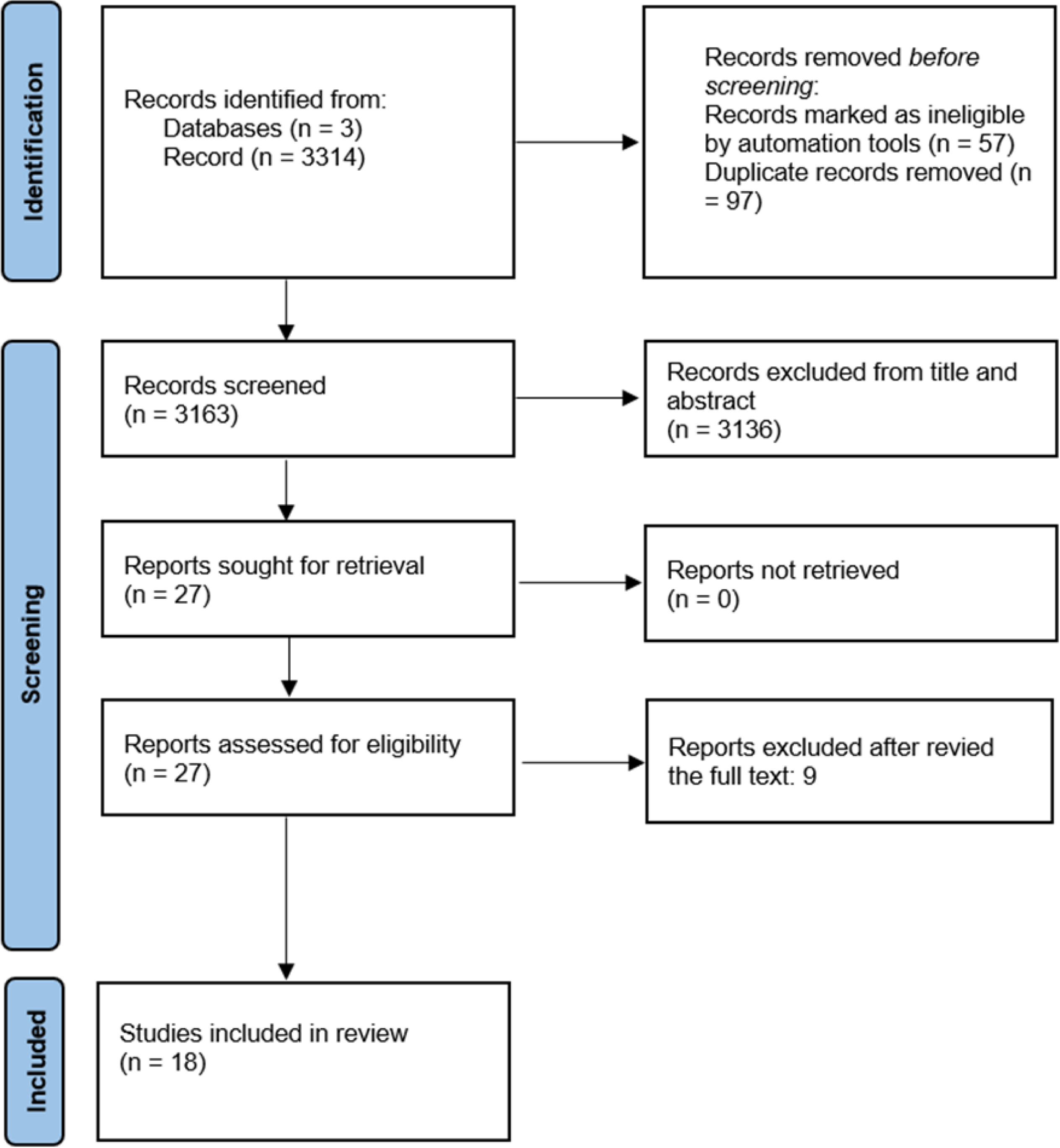
Flowchart of the mechanical thrombectomy studies subgroup.

Concerning PCI, the systematic review by Lun et al. included 133 studies(15). However, this study considered both acute and elective cases. To reduce clinical imbalance, only acute PCI setting were retrieved and included (33/133 studies [24.8%]).

The study characteristics and risk of bias assessments for both MT and PCI are summarized in **Tables 1 and S1**.

### Clinical results

A total of 69,464 patients were included in the analysis, including 30,138 (43.4%) who underwent MT and 39,326 (56.6%) who underwent PCI. The mean age of the patients included in the articles was 63.6 years ± 5.8, with an average percentage of male 66.6% ± 12.8. Only 31/51 (60.8%) papers specified a prevalence of preexisting chronic kidney disease (CKD) from 2.0% to 50.3%. A total of 45/51 (88.2%) articles reported a diabetes prevalence ranging from 11.1% to 53.0%. 46/51 (90.2%) studies reported smoking status, with a prevalence ranging from 7.5% to 72.0%.

The meta-analysis demonstrated high heterogeneity with I^2 98.32 and Cochrane’s Q 2984.77.

The AKI incidence in the entire population was 12.4% (9,390/69,464) (confidence interval (CI95%) 10.6 to 14.3]. The incidence of AKI in the MT subgroup was significantly lower compared to PCI subgroup (8.3% [CI95% 4.7 to 11.9] vs 14.7 [CI95% 12.6 to 16.8]; p<0.05) (**Figure 2a**.). The difference remained significant after removing the study by Alqahtani et al.(18) (7.8% [CI95% 5.1 to 10.5] vs 14.7% [CI95% 12.6 to 16.8], respectively).

**Figure 2.**
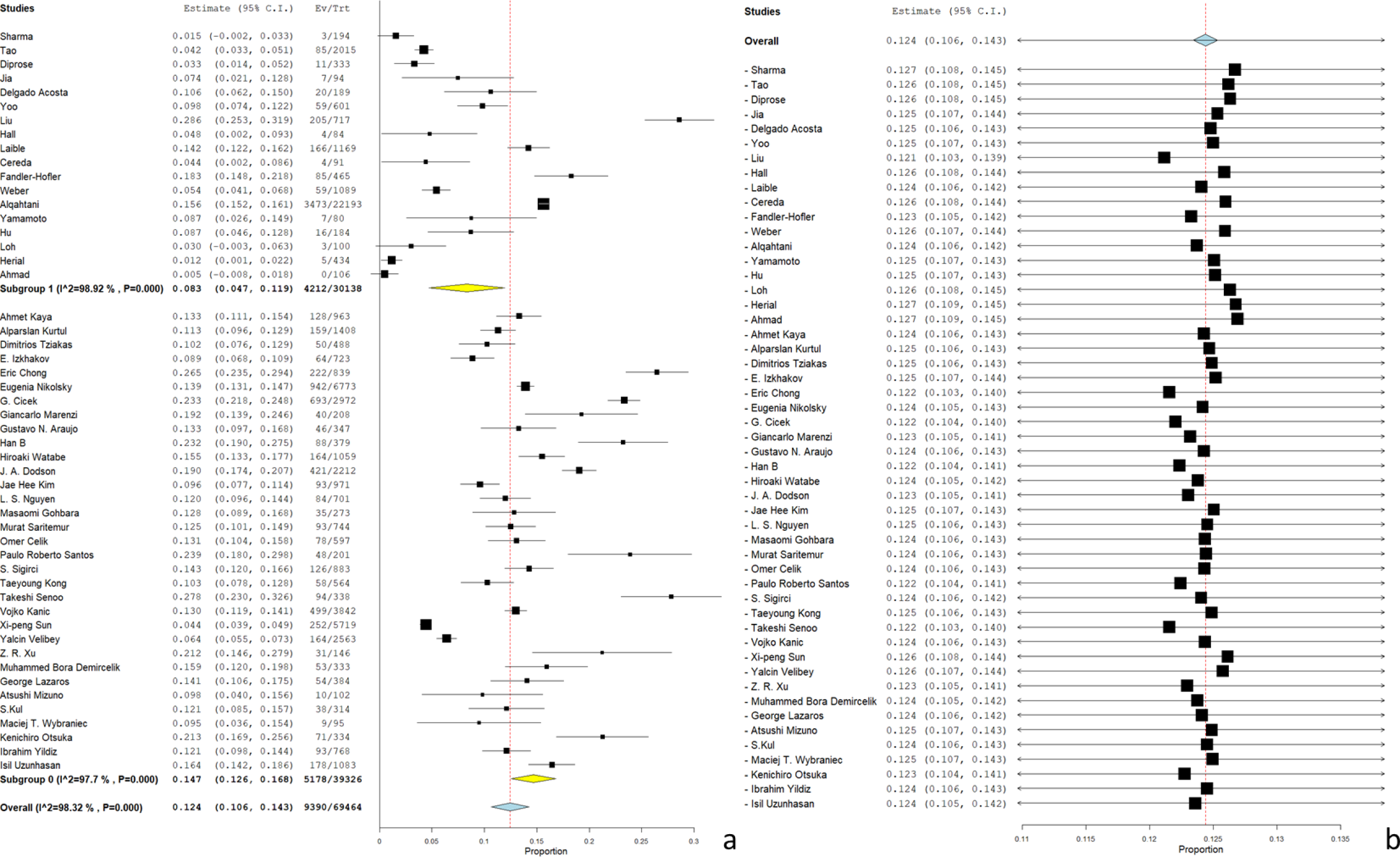
a) showed the meta-regression subgroups analysis; b) displayed the leaving-one-out meta-analysis.

### Sub-analysis on contrast media and IMA severity

The millilitres of contrast media administrated during PCI was significantly higher comparing with MT (198.6 [CI95% 178.1 to 219.30] vs 120.5 [CI95% 99.2 to 141.7]). Only 16/51 (31.4%) studies reported the use of a single contrast medium during all procedures with a range from 300 to 370 mg/ml; therefore, a comparison using iodine concentration as a covariate was not possible. The Killip classification was reported in 14/33 (42.4%) PIC studies, with a reported prevalence of Killip>1 of 11.3% (95% CI 7.5 14.7). The ejection fraction (EF) was included in 25/33 (75.8%) studies, with a mean of 49.1% ± 4.0.

### Meta-regression

In univariate analysis, only sex and smoking status were significantly associated with an increased risk of developing AKI (OR 1.002 [CI95% 1.001–1.004]; p=0.001 and OR 1.001 [CI95% 1.000–1.002]; p=0.033, respectively). In multivariable meta-regression analysis, CSIL was significantly associated with a decreased incidence of AKI (OR=0.93 [1.001–1.16]; p=0.03). A detailed analysis is presented in **Table 2**.

**Table 2.** Univariate and multivariate meta-regression for the prevalence of AKI. AKI acute kidney injury OR: odds ratio; CKD chronic kidney disease.

| Univariate |  |  |  |  | Multivariate |  |  |  |
| --- | --- | --- | --- | --- | --- | --- | --- | --- |
|  | OR | Lower Bound | Upper Bound | p | OR | Lower Bound | Upper Bound | p |
| Age | 1.001 | 0.997 | 1.004 | 0.61 |  |  |  |  |
| Gender (male) | 1.002 | 1.001 | 1.004 | 0.001 | 1.00 | 0.99 | 1.002 | 0.97 |
| Smoke | 1.001 | 1.000 | 1.002 | 0.033 | 1.00 | 0.999 | 1.002 | 0.90 |
| Diabetes | 1.002 | 1.000 | 1.005 | 0.06 |  |  |  |  |
| Hypertension | 1.000 | 0.998 | 1.001 | 0.733 |  |  |  |  |
| Contrast (ml) | 1.000 | 1.000 | 1.001 | 0.33 |  |  |  |  |
| Saline flush |  |  |  |  |  |  |  |  |
| CKD | 1.000 | 0.997 | 1.002 | 0.764 |  |  |  |  |
| No-Saline flush |  |  |  |  | 1.08 | 1.01 | 1.16 | 0.03 |

### Sensitive analysis

Leave-one-out meta-analysis did not reveal any significant differences (**Figure 2b**). Furthermore, the OR remained similar after removing Cereda et al.[19] (OR 1.08 [CI95% 1.01 to 1.16]), who derived data from an RCT (DEFUSE 3), and Alqahtani et al.(18), who impacted >70T of the sample size in the MT subgroup(OR 1.08 [CI95% 1.01 to 1.16]). Funnel plots showed asymmetry, which demonstrated moderate publication bias (**Figure 3**).

**Figure 3.**
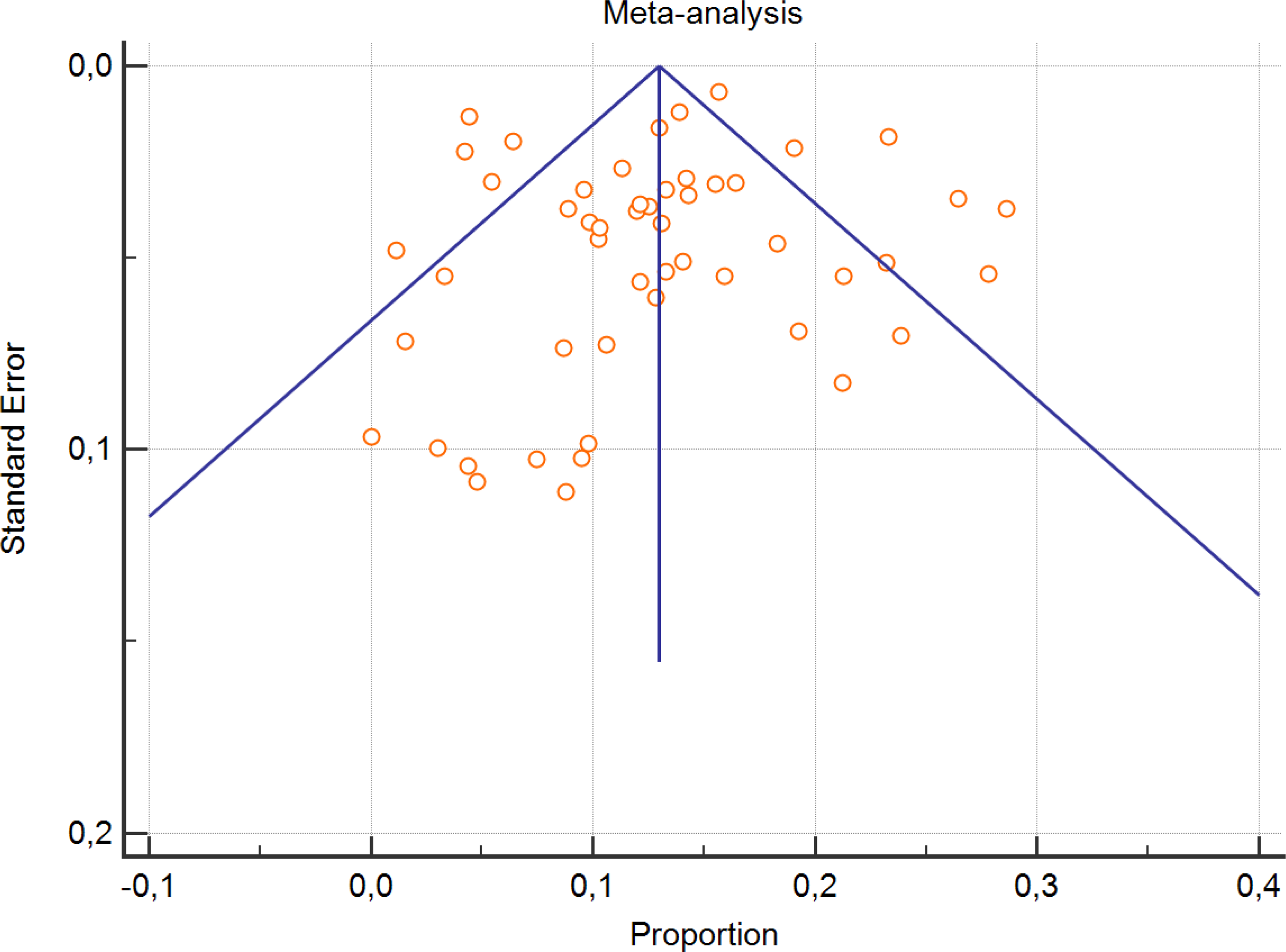
Funnel plot.

## Perspective

The present study demonstrated that CSIL during endovascular procedures (MT and PCI) may significantly and independently reduce the incidence of AKI in an emergency setting, including IS and AMI.

AKI is a major problem during hospitalization, accounting for an incidence of 10-15% for all hospitalizations, which increases to 50% in ICU patients(2). The incidence of AKI in both AMI and IS is similar (7.9 for PCI vs. 5.0% for MT (19,20)). AKI may occur for several reasons, including clinical conditions, drugs, and contrast medium administration(2). The occurrence of AKI during myocardial infarction increased the chance of mortality or major cardiovascular events, with a hazard ratio of 7.9, (95% CI 2.5 to 13)(21).

The pathophysiology of AKI depends on conditions associated with its develop(2). Concerning the development of AKI during stroke or myocardial infarction treated with endovascular procedures (MT and PCI, respectively), the pathophysiology is similar, and includes intermittent hypovolemia, inflammation, and contrast medium administration(22,23). In addition, a cardiac pump deficit after AMI may play a role in the development of AKI by perturbations in the renal blood flow, leading to an imbalance in oxygen supply and demand(2). Unfortunately, no MT studies in this meta-analysis reported the ejection fraction (EF%) or heart failure score systems (e.g., Killip classification). However, the pooled absolute value of EF%, in the PCI subgroups, was only slightly inferior comparing with normality (49.1% ± 4.0)(24). Furthermore, the prevalence of Killip score >1 is inferior to that reported in the literature (11.3% vs. 33.2%) after AMI (25). In addition, cardiovascular damage is the second leading cause of death after stroke, and 28.5% of patients had LVEF < 50%, and 13–29% had systolic dysfunction at 3 months(26). This evidence suggests that although a direct comparison regarding cardiac pump deficit is not possible due to lack of data, it can be assumed that both populations are similar in this respect.

Hydration plays a key role in the prevention and treatment of AKI (Level B evidence)(13). CSIL is a standard operative procedure during MT to reduce the risk of thrombus formation inside the catheters(8). The idea behind this study is that CSIL, which has the secondary effect of fluid expander, during MT is responsible for the lower incidence of AKI compared with that after PCI in AMI. To demonstrate this, we performed a subgroup meta-analysis comparing the incidence of AKI in both the MT and PTCA data subsets (MT 8.3% [CI95% 4.7 to 11.9] vs PCI 14.7 [CI95% 12.6 to 16.8]) (**Figure 2a. and 2b.**). These data were similar to those extracted from the literature for IS (5.0% [95% CI, 2.1% to 8.9%])(20) and AMI (12.8% [95% CI 11.7 to 13.9%]) (15).

Although acute myocardial infarction and ischemic stroke share a clinical picture, prognosis, and effective treatment, some differences exist(5). However, the admission rates in intensive care and short-term mortality were similar between IS and AMI (88% vs. 82.3% and 15% vs. 10%, respectively), showing a similar clinical course(27–30). In this systematic review, the amount of contrast media injected intra-arterial was significantly higher during PCI comparing with MT (198.6 [CI95% 178.1 to 219.30] vs 120.5 [CI95% 99.2 to 141.7]). However, the meta-regression did not demonstrate a significant correlation between AKI and the amount of contrast medium injected (OR 1 [CI95 1-1.001], p=0.33).

Nevertheless, to balance the clinical conditions between the two diseases, meta-regression using clinical/demographic details as covariates was performed (**Table 2**). As hypostatized in the multivariate analysis, the absence of CSIL was associated with a significant increase in the risk of developing AKI (OR 1.08 [CI95% 1.001–1.16]; p = 0.03).

However, all MT use CSIL and, conversely, all PCI do not; therefore, a multicollinearity problem is evident. However, the results have a strong pathophysiological meaning: CSIL is a fluid expander, which is the treatment/prevention strategy for AKI(31,32).

In fact, from a scientific/clinical point of view, since hydration using saline is the standard strategy for treatment and preventing AKI, CSIL during MT may have the same effect (13,33,34). Although it is not possible to determine the exact amount of saline administered during the procedure, it can be estimated in a range of 361.8 to 1085.4 ml for a mean procedural time of 67 min and 3 catheter lines(10,11). The estimated amount of saline administered during MT is similar to that suggested by the ESUR Contrast Medium Safety Committee guidelines (800 ml for a patient weighing 80 kg)(13). This hypothesis is also indirectly supported by the lower incidence of AKI depicted between patients treated with MT and all ischemic stroke (1.6% compared with 4.2%; p = 0.0071)(35). This is even more important considering that patients treated with MT tend to have more severe clinical conditions and more contrast media administration (also intra-arterial), and therefore, a higher theoretical expected AKI incidence(6). However, the incidence is significantly lower (relative risk MT 0.4 [95% CI 0.2 to 0.8], p<0.007) (35). On the contrary, the administration of fibrinolytic therapy (rt-PA) increase the chance of having AKI (relative risk 1.9 [95% CI 1.1 to 3.3], p<0.03)(35). A potential explanation for this apparent discrepancy is the CSIL during MT. Furthermore, this lower incidence of AKI reported for MT is not observed in acute myocardial infarction treated or not treated endovascularly (with PCI 8.6% versus without PCI 10.9%, p=0.12 and OR 0.79 [95% CI 0.60 to 1.05], p=0.10)(36). Furthermore, after propensity score one-on-one match, the incidence between PCI and no-PCI patients is similar (8.6% vs 10.9%, OR 0.77 [95% CI, 0.56–1.06] p=0.12)(36).

The potential reduction in the incidence of AKI during PCI for AMI using a simple method such as CSIL has relevant clinical implications. In fact, the occurrence of AKI during AMI increases the chance of death with an OR of 12.52 (95% CI 9.29–16.86)(37). In addition, for nine cases of AKI successfully avoided during AMI treated with PCI, one death could be prevent(37).

This study has several limitations. First, although the results were consistent with the pathophysiologywasevention, and treatment theof AKI, multicollinearity remains a potential source of bia.sAlthough, a direct comparison regarding cardiac pump deficits were not possible, although, literature permits the assumption of clinical similarities although the results were consistent with the pathophysiology, prevention, and treatment of AKI, multicollinearity remains a potential source of bias. Third, ecological bias is an unresolved issue in meta-regressions. Fourth, we did not re-perform the meta-analysis by Lun et al.(15) concerning the incidence of AKI in PCI; however, the study had a large sample size (>1.2 million patients) and updated the data (published in 2021).

In conclusion, CSIL significantly reduced the incidence of AKI during endovascular procedures in the emergent setting including ischemic stroke and acute myocardial infarction. Due to the limitations of the present review, a randomized controlled trial that compared the efficacy of CSIL in PCI is mandatory owing to its clinical implications.

## Data Availability

The data are published available being a systematic review

## Abbreviation list

AKI: acute kidney injury
PCI: percutaneous coronary intervention
MT: mechanical thrombectomy
IS: ischemic stroke
AMI: acute myocardial infarction
CSIL: continuous saline infusion line
OR: odds ratio

